# A Bayesian Survival Analysis on Long COVID and non Long COVID patients: A Cohort Study Using National COVID Cohort Collaborative (N3C) Data

**DOI:** 10.1101/2024.06.25.24309478

**Authors:** Sihang Jiang, Johanna Loomba, Andrea Zhou, Suchetha Sharma, Saurav Sengupta, Jiebei Liu, Donald Brown, N3C consortium

## Abstract

Since the outbreak of COVID-19 pandemic in 2020, numerous researches and studies have focused on the long-term effects of COVID infection. The Centers for Disease Control (CDC) implemented an additional code into the International Classification of Diseases, Tenth Revision, Clinical Modification (ICD-10-CM) for reporting ‘Post COVID-19 condition, unspecified (U09.9)’ effective on October 1st 2021, representing that Long COVID is a real illness with potential chronic conditions. The National COVID Cohort Collaborative (N3C) provides researchers with abundant electronic health records (EHR) data by aggregating and harmonizing EHR data across different clinical organizations in the United States, making it convenient to build up a survival analysis on Long COVID patients and non Long COVID patients among large amounts of COVID positive patients.

## 1 Introduction

The outbreak of COVID-19 pandemic since 2020 has impacted everyone. As time goes by, it is of more interest to focus on the long-term effect of COVID-19. According to Centers for Disease Control and Prevention (CDC), Long COVID is broadly defined as signs, symptoms, and conditions that continue or develop after acute COVID-19 infection [1]. Some conditions can last weeks, months, or years. The diagnosis code (U09.9) for Long COVID implemented by CDC [2] makes it much easier to identify this disease.

With stewardship from National Center for Advancing Translational Sciences (NCATS) and data contributions from more than 75 institutions, the National COVID Cohort Collaborative (N3C) [3] is one of the largest collections of clinical data related to COVID-19 patients in the United States, including more than 7 million COVID positive patients and more than 20 billion rows of electronic health records (EHR) data for cohort studies. Since the implementation of U09.9 code, various researches have focused on the characteristics of Long COVID using machine learning methods on different cohorts, including risk factors, subtypes and vital measurements [4] [5] [6] [7], and efforts have been put into understanding Long COVID better.

## 2 Related Work

Two main categories of biases in cohort studies include selection bias and information bias [8]. In a cohort study of COVID patients, two elements of selection bias, sampling bias and confounding by indication, might lead to an unrepresentative sample of the population; and among information bias, observer bias and lead-time bias on Long COVID patients might affect the accuracy of survival analysis.

Competing risk in survival analysis is common. A competing risk is an event whose occurrence precludes the occurrence of the primary event of interest. In studies on cardiovascular disease and depression respectively [9] [10], failure to account correctly for competing events can result in unexpected consequences, including overestimation of the probability of the event and mis-estimation of the magnitude of relative effects of covariates on the outcome.

According to CDC [1], Since October 2021, Long COVID is a real illness and can result in chronic conditions including respiratory and heart symptoms, neurological symptoms, digestive symptoms and so on. For some people, these symptoms can last weeks, months, or years. In a recent study on the risk factors of Long COVID [4], it is shown that middle-age (40 to 69 years), female sex, hospitalization associated with COVID-19, long hospitalization stay, receipt of mechanical ventilation, and several comorbidities including depression, chronic lung disease, and obesity are associated with higher likelihood of Long COVID.

There are various studies on survival of COVID patients since the pandemic. A research on Brazilian COVID patients showed that old age and cardiovascular disease are associated with higher mortality [11], and a study on COVID patients admitted to intensive care unit showed heterogeneity in the survival [12]. Another study on COVID and kidney diseases mentioned that acute kidney disease and chronic kidney disease are associated with higher risk of death, and COVID might lead to chronic kidney disease in survivors [13].

After the release of U09.9 [2] code of Long COVID, it is of great interest to study characteristics of Long COVID patients, but there are limited researches on the survival analysis, and the comparison between Long COVID patients and non Long COVID patients. This paper applies a Bayesian survival analysis approach on Long COVID patients and non Long COVID patients, and study the features associated with mortality.

## 3 Methodology

### 3.1 Survival Analysis

Survival analysis mainly focus on time-to-event data. Following the notation in [14], we introduce several important variables and functions in survival analysis under Bayesian paradigm. Let *T* be a continuous nonnegative random variable representing the survival times of individuals in a population, defined over the interval [0, ∞). Let *f* (*t*) denote the probability density function (pdf) of *T*, and the cumulative distribution function (cdf) of *T* is

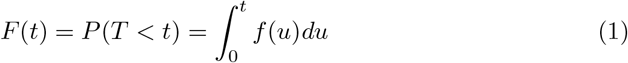

and the survivor function to describe the probability of surviving till time *t* is

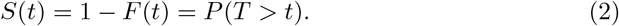

The hazard function *h*(*t*), the instant rate of failure at time *t* is defined as

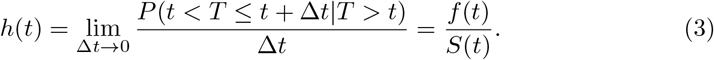

Censoring is common in survival data, and it occurs when incomplete information is available about the survival time of some individuals [15]. An observation is said to be right censored at *c* if the exact value of the observation is not known but only that it is greater than or equal to *c*; an observation is said to be left censored at *c* if it is known only that the observation is less than or equal to *c*; an observation is said to be interval censored if it is known only that the observation is in the interval (*c*_1_, *c*_2_). Type-I censoring and Type-II censoring [16] [17] are commonly used in different parametric models as well in survival analysis.

### 3.2 Bayesian Parametric Models

Under Bayesian paradigm [14], given unknown parameters we first set up a prior distribution, and combine the likelihood function with the prior distribution to get the posterior distribution of parameters. Markov chain Monte Carlo (MCMC) methods are widely used in sampling from a complicated distribution, such as Gibbs sampling, Metropolis-Hasting algorithm, and Hamiltonian Monte Carlo [18] [19]. PyMC [20] is a Python module allowing users to implement Bayesian statistical models with different parameters, prior distributions and likelihood functions, as well as calculating the numerical results of posterior estimation of parameters.

Suppose we have independent identically distributed (i.i.d.) survival time ***y*** = (*y*_1_, …, *y*_*n*_)^*T*^ with right censor indicator ***ν*** = (*ν*_1_, …, *ν*_*n*_)^*T*^ where *ν*_*i*_ = 1 if *y*_*i*_ is an observed failure time, and *ν*_*i*_ = 0 if *y*_*i*_ is right censored. Let *D* = (*n*, ***y, ν***), we consider the following models.

#### 3.2.1 Log-normal Model

The log-normal model is a two-parameter model. The survival time *y*_*i*_ has a log-normal distribution defined on (0, +∞) with density function, mean and variance and survival function

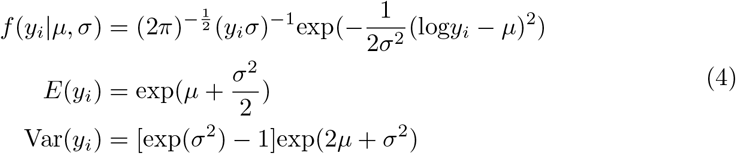

and

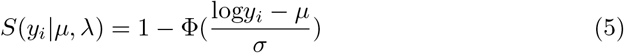

with Φ(.) representing the cdf of the standard normal distribution. The likelihood is

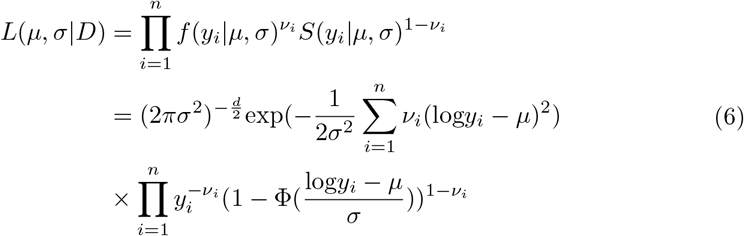

Let *τ* = 1*/σ*^2^, and a common prior distribution *p*(*μ, τ*) assumes a normal distribution on *μ* and a gamma distribution on *τ*. The posterior distribution is given by

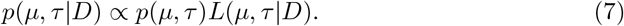

To build a regression model, we introduce covariates through *μ* and write 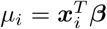. Common prior distributions of ***β*** include uniform improper prior and normal prior.

## 4 Data and Results

### 4.1 Cohort Identification

In this study, the cohort consists of a group of Long COVID patients and a group of non Long COVID patients. Patients of both groups have evidence of COVID-19. According to CDC, Long COVID starts to be identified after several weeks of COVID infection [1], leading to possible selection bias in the cohort while conducting survival analysis. Thus, we implement appropriate inclusion criteria to control the selection bias. In addition, among all COVID positive patients in N3C, only a very small number of patients are diagnosed with Long COVID by U09.9 code. To make a balanced cohort, we use 1-on-1 matching [21], assigning each Long COVID patient to a COVID positive control.

#### 4.1.1 Long COVID group inclusion criteria

- COVID-19 positive patients as defined with a positive SARS-CoV-2 PCR/AG test or a recorded U07.1 diagnosis. The earliest date of either will be their index date.
- Patients with a U09.9 code.
- The U09.9 diagnosis code should be no earlier than Oct. 1st 2021.
- The U09.9 diagnosis code should be no earlier than the COVID index date.
- Patients should be greater or equal to 18 years old.

#### 4.1.2 COVID positive control group inclusion criteria

- COVID-19 positive patients as defined with a positive SARS-CoV-2 PCR/AG test or a recorded U07.1 positive diagnosis. The earliest date of either will be their index date.
- Patients should be greater or equal to 18 years old.
- Patients with no U09.9 code
- Selected based on the matching process to ensure a 1-to-1 ratio to Long COVID group

#### 4.1.3 Matching process

To make the comparison between the Long COVID group and the control group more reliable, we try to select patients with similar health conditions in the matching process, using variables including age, health system site id, COVID index date, number of clinical visits and Charlson comorbidity index (CCI) [22].

The matching process will be done without replacement, and the criteria for matching each pair of Long COVID patient and COVID positive control patient are as following:

- The Long COVID patient and the COVID positive control are from the same health system.
- The age difference between the Long COVID patient and the COVID positive control are less than or equal to 10 years.
- The difference of COVID index date between the Long COVID patient and the COVID positive control are less than or equal to 45 days.
- The difference of the log of number of visits before COVID index date between the Long COVID patient and the COVID positive control are less than or equal to 1.
- The difference of the log of CCI score through COVID index date between the Long COVID patient and the COVID positive control are less than or equal to 0.5.

In this survival analysis, we choose the U09.9 code date for any matched pair as day 0 of survival. The death date is from a combination of OMOP death records [23] and PPRL death records [24] in N3C. We drop a pair of patients if anyone in this pair has a death date earlier than U09.9 code date. The survival length is the outcome of the survival analysis, defined as the time difference between day 0 of survival and the earlier of death date and April 1st 2024, as completion of this study.

### 4.2 Cohort Summary

As the completion of the study, there are 7,376,162 COVID positive patients in N3C as in the COVID summary table, and among them there are only 74,240 patients with the U09.9 diagnosis code. Cohort characteristics before the matching process and after the matching process are both included below. Table 1 and Table 2 are the attrition tables before the matching process, and Table 3 and Table 4 show characteristics of patients before the matching process and the final cohort.

**Table 1.** Long COVID group attrition table before matching.

| inclusion criteria | number of patients |
| --- | --- |
| COVID positive patients in N3C | 7,376,162 |
| patients with U09.9 code | 74,240 |
| U09.9 date later than COVID index date and 2021-10-01 | 72,121 |
| age greater or equal to 18 | 69,805 |

**Table 2.** COVID positive control group attrition table before matching.

| inclusion criteria | number of patients |
| --- | --- |
| COVID positive patients in N3C | 7,376,162 |
| age greater or equal to 18 | 6,202,631 |
| patients with no U09.9 code | 6,127,387 |
| patients from clinical sites reporting U09.9 | 5,431,788 |

**Table 3.** Summary table before matching.

|  | COVID positive control group | Long COVID group |
| --- | --- | --- |
| number of patients | 5,431,788 | 69,805 |
| age (mean(SD)) | 47.92 (18.79) | 54.84 (16.50) |
| sex (%) |  |  |
| Female | 3,070,884 (56.5) | 45,314 (64.9) |
| Male | 2,340,669 (43.1) | 24,038 (34.4) |
| race and ethnicity (%) |  |  |
| Asian Non-Hispanic | 152,340 (2.8) | 1,853 (2.7) |
| Black or African American Non-Hispanic | 660,480 (12.2) | 8,014 (11.5) |
| White Non-Hispanic | 3,452,553 (63.6) | 48,898 (70.0) |
| Other Non-Hispanic | 205,047 (3.8) | 1,473 (2.1) |
| Hispanic or Latino Any Race | 628,286 (11.6) | 6,656 (9.5) |
| Missing/Unknown | 333,082 (6.1) | 2,911 (4.2) |
| CCI category (%) |  |  |
| 0 | 3,653,286 (67.3) | 28,598 (41.0) |
| 1-2 | 1,066,217 (19.6) | 22,129 (31.7) |
| 3-4 | 378,595 (7.0) | 9,757 (14.0) |
| 4+ | 333,690 (6.1) | 9,321 (13.4) |
| age category (%) |  |  |
| 18-20 | 263,208 (4.8) | 850 (1.2) |
| 21-45 | 2,350,131 (43.3) | 20,193 (28.9) |
| 46-65 | 1,724,495 (31.7) | 29,414 (42.1) |
| 66+ | 1,093,954 (20.1) | 19,348 (27.7) |

**Table 4.**
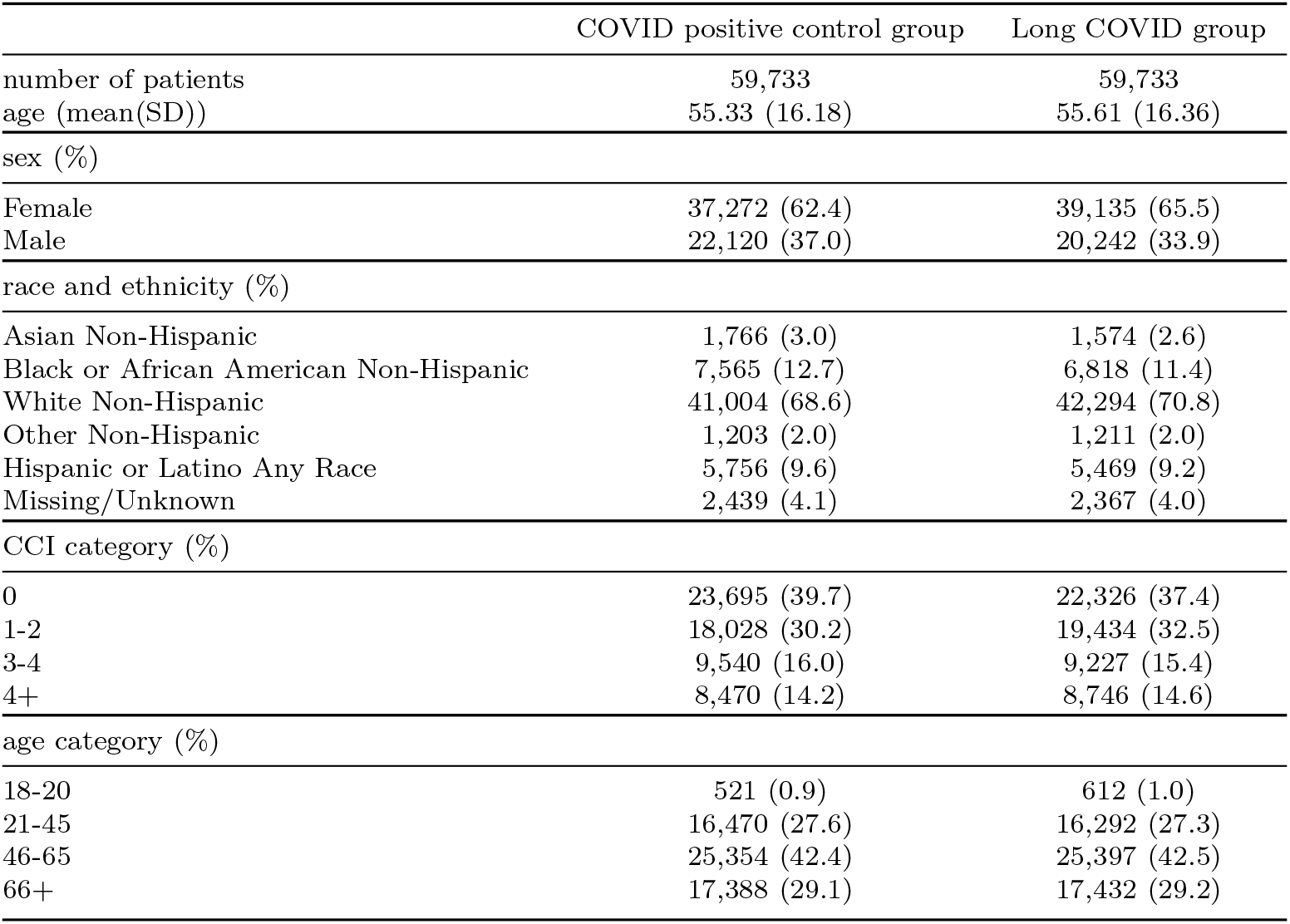
Final cohort summary table.

After the matching process, the final cohort has 119,466 patients in total with 59,733 Long COVID (U09.9) patients and 59,733 COVID positive control patients.

### 4.3 Modeling

Following the notations in 3, *y*_*i*_ is the survival time of a patient in the cohort, with the distribution

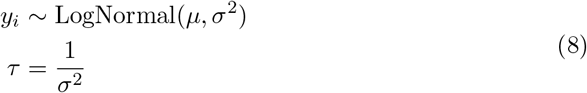

where

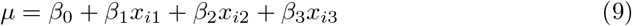

and *x*_*i*1_, *x*_*i*2_, *x*_*i*3_ represent 3 features in the model: whether the patient has Long COVID (U09.9), whether the patient has obesity (BMI greater or equal to 30 [25]) and whether the COVID symptom of the patient is mild (mild COVID is defined as no emergency department visit nor hospitalization around COVID index date). The prior distributions of the parameters are as following:

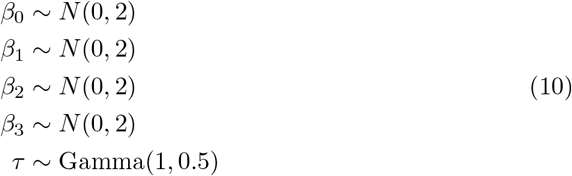

### 4.4 Results

Given prior distributions and likelihood specified b y t he m odeling, t he p osterior of parameters are numerically estimated using Markov Chain Monte Carlo (MCMC) in Pymc [20]. Table 5 shows the posterior mean, standard deviation and 95% high-density interval of parameters, and Fig. 1 shows the trace plots of parameters in the MCMC sampling process.

**Table 5.** Posterior estimation of parameters.

| parameter | mean | standard deviation | 95% High-Density Interval |
| --- | --- | --- | --- |
| $\beta_0$ | 6.146 | 0.006 | [6.135, 6.157] |
| $\beta_1$ | -0.009 | 0.005 | [-0.018, 0] |
| $\beta_2$ | 0.005 | 0.005 | [-0.004, 0.014] |
| $\beta_3$ | 0.094 | 0.005 | [0.084, 0.104] |
| $\tau$ | 1.754 | 0.008 | [1.739, 1.769] |

**Fig. 1.**
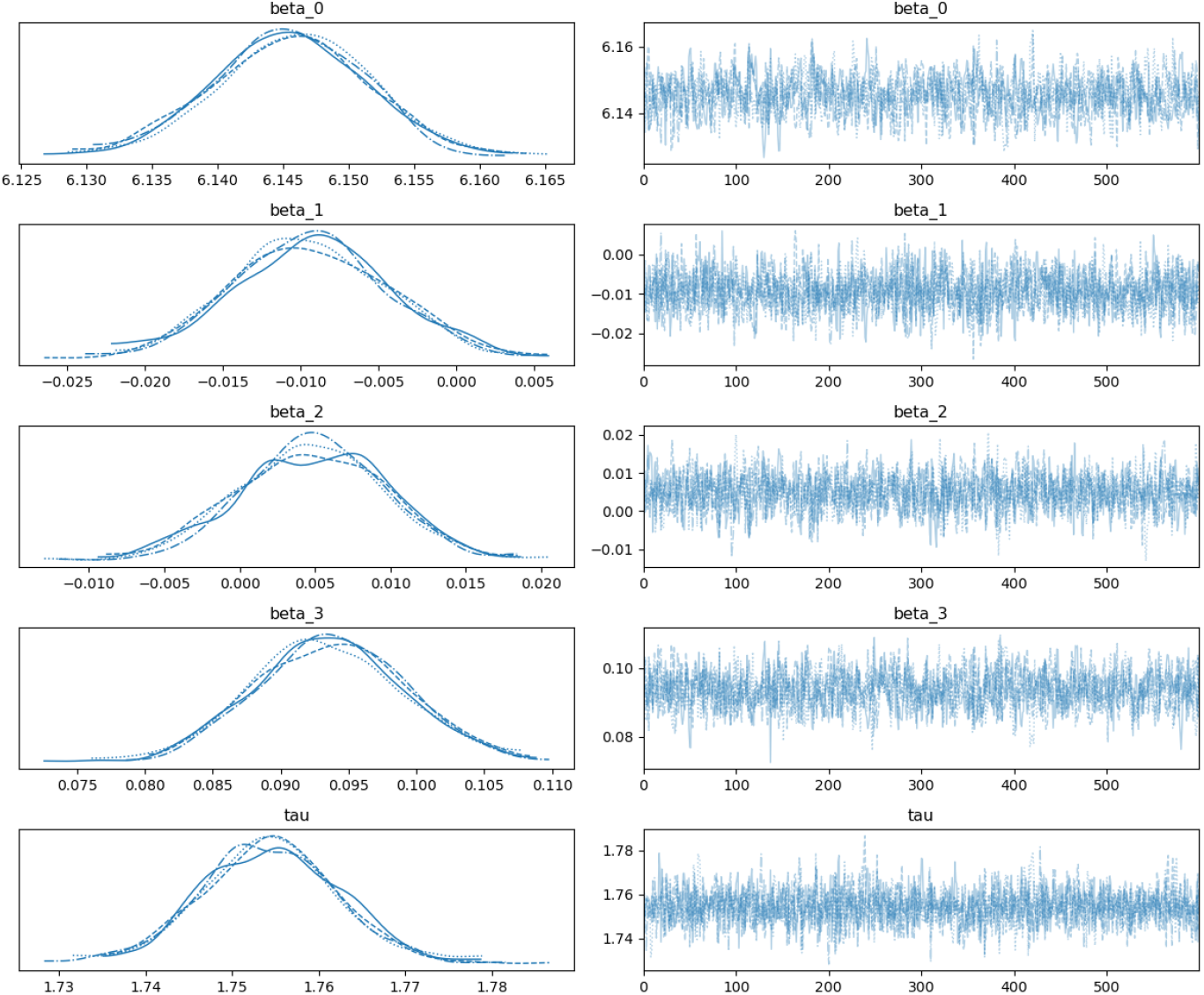
Posterior estimations of parameters by MCMC

## 5 Conclusion

According to the posterior estimation of parameters, patients with Long COVID (U09.9) indicator are more likely to have shorter survival length, patients with mild COVID symptoms are more likely to have longer survival length, and the obesity indicator is not significant with respect to the survival length.

The Kaplan-Meier estimator is a non-parametric method to estimate the survival probability from lifetime data [26]. Fig. 2 shows the Kaplan-Meier curves of two groups in the cohort, and according to the curves, the Long COVID group has lower survival probability than the COVID positive control group from day 0 of survival til the maximum observed survival length.

**Fig. 2.**
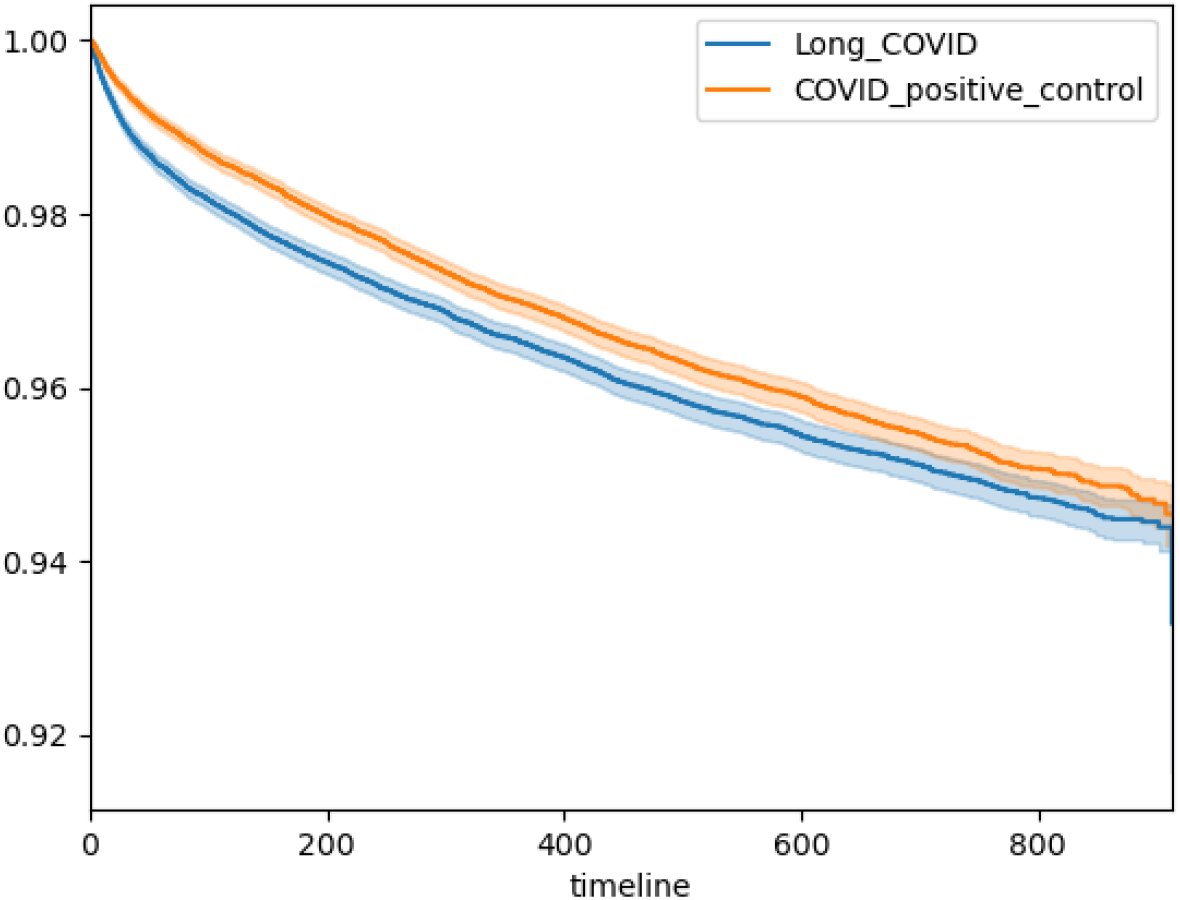
Kaplan-Meier curves of two groups in the cohort

## 6 Limitation and Future Work

Since the CDC issued U09.9 code in October 2021, the maximum possible survival length of a Long COVID patient is less than 1,000 days. According to Fig. 3, the distribution of survival length is left skewed. This paper uses a parametric method [14], assuming the observations of survival length are from a log-normal distribution. Other distributions should be taken into consideration in order to accommodate the left skewness, such as Weibull distribution and Gamma distribution.

**Fig. 3.**
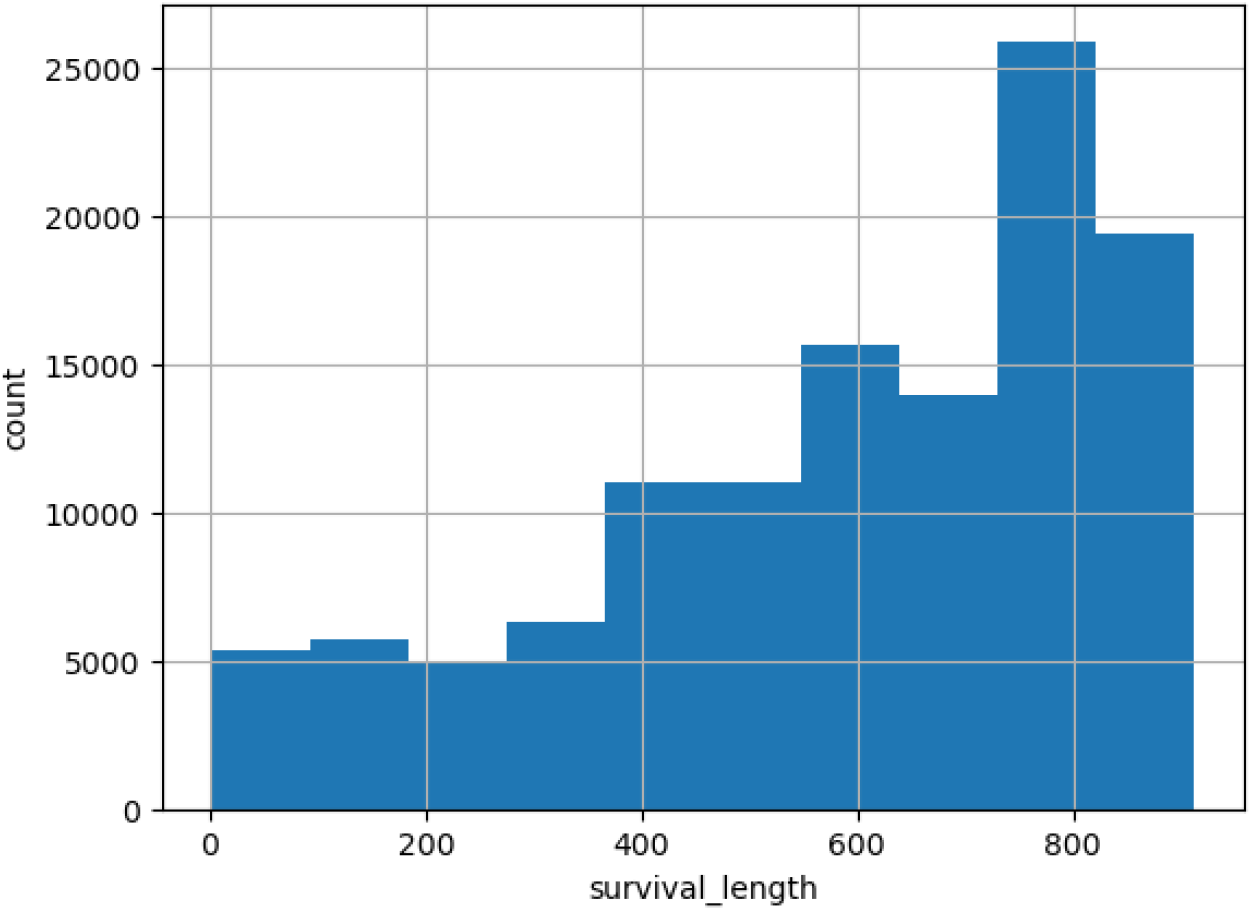
Survival length of the cohort

In a parametric model, the cumulative distribution function of the survival length is assumed to be differentiable, but this assumption might not hold. Semiparametric models mainly focus on the baseline hazard or cumulative hazard, and a common example is the piecewise constant hazard model, where the hazard rate might differ in time periods, and different subgroups [27] [28].

Among all COVID patients in N3C, Long COVID (U09.9) patients are a very small portion. The purpose of the matching process is creating a balanced cohort as well as controlling the selection bias [8]. However, there are limited discussions on the features to use in the matching process, and how the matching process will affect the accuracy of modeling. The possibility of new bias created from the matching process cannot be ignored.

There are few studies on the censoring of Long COVID patients. In this paper, the observations of survival length are assumed to be not censored for convenience of calculation. Whether there exists censoring and what type censoring in survival length are still under question in study of Long COVID patients [29] [30].

## Data Availability

All data produced in the present study are available upon reasonable request to the authors

## N3C Attribution

The analyses described in this publication were conducted with data or tools accessed through the NCATS N3C Data Enclave https://covid.cd2h.org and N3C Attribution & Publication Policy v 1.2-2020-08-25b supported by NCATS Contract No. 75N95023D00001, Axle Informatics Subcontract: NCATS-P00438-B, and iTHRIV Integrated Translational health Research Institute of Virginia UL1TR003015. This research was possible because of the patients whose information is included within the data and the organizations (https://ncats.nih.gov/n3c/resources/data-contribution/data-transfer-agreement-signatories) and scientists who have contributed to the on-going development of this community resource [3].

## Disclaimer

The N3C Publication committee confirmed that this manuscript MSID:1996.791 is in accordance with N3C data use and attribution policies; however, this content is solely the responsibility of the authors and does not necessarily represent the official views of the National Institutes of Health or the N3C program.

## IRB

The N3C data transfer to NCATS is performed under a Johns Hopkins University Reliance Protocol # IRB00249128 or individual site agreements with NIH. The N3C Data Enclave is managed under the authority of the NIH; information can be found at https://ncats.nih.gov/n3c/resources.

## Individual Acknowledgements For Core Contributors

We gratefully acknowledge the following core contributors to N3C:

Adam B. Wilcox, Adam M. Lee, Alexis Graves, Alfred (Jerrod) Anzalone, Amin Manna, Amit Saha, Amy Olex, Andrea Zhou, Andrew E. Williams, Andrew Southerland, Andrew T. Girvin, Anita Walden, Anjali A. Sharathkumar, Benjamin Amor, Benjamin Bates, Brian Hendricks, Brijesh Patel, Caleb Alexander, Carolyn Bramante, Cavin Ward-Caviness, Charisse Madlock-Brown, Christine Suver, Christopher Chute, Christopher Dillon, Chunlei Wu, Clare Schmitt, Cliff Takemoto, Dan Housman, Davera Gabriel, David A. Eichmann, Diego Mazzotti, Don Brown, Eilis Boudreau, Elaine Hill, Emily Carlson Marti, Emily R. Pfaff, Evan French, Farrukh M Koraishy, Federico Mariona, Fred Prior, George Sokos, Greg Martin, Harold Lehmann, Heidi Spratt, Hemalkumar Mehta, J.W. Awori Hayanga, Jami Pincavitch, Jaylyn Clark, Jeremy Richard Harper, Jessica Islam, Jin Ge, Joel Gagnier, Johanna Loomba, John Buse, Jomol Mathew, Joni L. Rutter, Julie A. McMurry, Justin Guinney, Justin Starren, Karen Crowley, Katie Rebecca Bradwell, Kellie M. Walters, Ken Wilkins, Kenneth R. Gersing, Kenrick Dwain Cato, Kimberly Murray, Kristin Kostka, Lavance Northington, Lee Allan Pyles,, Lesley Cottrell, Lili Portilla, Mariam Deacy, Mark M. Bissell, Marshall Clark, Mary Emmett, Matvey B. Palchuk, Melissa A. Haendel, Meredith Adams, Meredith Temple-O’Connor, Michael G. Kurilla, Michele Morris, Nasia Safdar, Nicole Garbarini, Noha Sharafeldin, Ofer Sadan, Patricia A. Francis, Penny Wung Burgoon, Philip R.O. Payne, Randeep Jawa, Rebecca Erwin-Cohen, Rena Patel, Richard A. Moffitt, Richard L. Zhu, Rishi Kamaleswaran, Robert Hurley, Robert T. Miller, Saiju Pyarajan, Sam G. Michael, Samuel Bozzette, Sandeep Mallipattu, Satyanarayana Vedula, Scott Chapman, Shawn T. O’Neil, Soko Setoguchi, Stephanie S. Hong, Steve Johnson, Tellen D. Bennett, Tiffany Callahan, Umit Topaloglu, Valery Gordon, Vignesh Subbian, Warren A. Kibbe, Wenndy Hernandez, Will Beasley, Will Cooper, William Hillegass, Xiaohan Tanner Zhang. Details of contributions available at covid.cd2h.org/core-contributors

## Data Partners with Released Data

The following institutions whose data is released or pending:

Available: Advocate Health Care Network — UL1TR002389: The Institute for Translational Medicine (ITM) · Aurora Health Care Inc — UL1TR002373: Wisconsin Network For Health Research · Boston University Medical Campus — UL1TR001430: Boston University Clinical and Translational Science Institute · Brown University — U54GM115677: Advance Clinical Translational Research (Advance-CTR) · Carilion Clinic — UL1TR003015: iTHRIV Integrated Translational health Research Institute of Virginia · Case Western Reserve University — UL1TR002548: The Clinical & Translational Science Collaborative of Cleveland (CTSC) · Charleston Area Medical Center — U54GM104942: West Virginia Clinical and Translational Science Institute (WVCTSI) · Children’s Hospital Colorado — UL1TR002535: Colorado Clinical and Translational Sciences Institute · Columbia University Irving Medical Center UL1TR001873: Irving Institute for Clinical and Translational Research · Dartmouth College — None (Voluntary) Duke University — UL1TR002553: Duke Clinical and Translational Science Institute · George Washington Children’s Research Institute — UL1TR001876: Clinical and Translational Science Institute at Children’s National (CTSA-CN) · George Washington University — UL1TR001876: Clinical and Translational Science Institute at Children’s National (CTSA-CN) · Harvard Medical School — UL1TR002541: Harvard Catalyst · Indiana University School of Medicine — UL1TR002529: Indiana Clinical and Translational Science Institute · Johns Hopkins University — UL1TR003098: Johns Hopkins Institute for Clinical and Translational Research · Louisiana Public Health Institute — None (Voluntary) · Loyola Medicine — Loyola University Medical Center · Loyola University Medical Center — UL1TR002389: The Institute for Translational Medicine (ITM) · Maine Medical Center — U54GM115516: Northern New England Clinical & Translational Research (NNE-CTR) Network · Mary Hitchcock Memorial Hospital & Dartmouth Hitchcock Clinic — None (Voluntary) · Massachusetts General Brigham — UL1TR002541: Harvard Catalyst · Mayo Clinic Rochester — UL1TR002377: Mayo Clinic Center for Clinical and Translational Science (CCaTS) · Medical University of South Carolina — UL1TR001450: South Carolina Clinical & Translational Research Institute (SCTR) · MITRE Corporation — None (Voluntary) · Montefiore Medical Center — UL1TR002556: Institute for Clinical and Translational Research at Einstein and Montefiore · Nemours — U54GM104941: Delaware CTR ACCEL Program · NorthShore University HealthSystem — UL1TR002389: The Institute for Translational Medicine (ITM) · Northwestern University at Chicago — UL1TR001422: Northwestern University Clinical and Translational Science Institute (NUCATS) · OCHIN — INV-018455: Bill and Melinda Gates Foundation grant to Sage Bionetworks · Oregon Health & Science University — UL1TR002369: Oregon Clinical and Translational Research Institute · Penn State Health Milton S. Hershey Medical Center — UL1TR002014: Penn State Clinical and Translational Science Institute · Rush University Medical Center — UL1TR002389: The Institute for Translational Medicine (ITM) · Rutgers, The State University of New Jersey — UL1TR003017: New Jersey Alliance for Clinical and Translational Science · Stony Brook University — U24TR002306 · The Alliance at the University of Puerto Rico, Medical Sciences Campus — U54GM133807: Hispanic Alliance for Clinical and Translational Research (The Alliance) · The Ohio State University — UL1TR002733: Center for Clinical and Translational Science · The State University of New York at Buffalo — UL1TR001412: Clinical and Translational Science Institute · The University of Chicago — UL1TR002389: The Institute for Translational Medicine (ITM) · The University of Iowa — UL1TR002537: Institute for Clinical and Translational Science · The University of Miami Leonard M. Miller School of Medicine — UL1TR002736: University of Miami Clinical and Translational Science Institute · The University of Michigan at Ann Arbor — UL1TR002240: Michigan Institute for Clinical and Health Research · The University of Texas Health Science Center at Houston — UL1TR003167: Center for Clinical and Translational Sciences (CCTS) · The University of Texas Medical Branch at Galveston — UL1TR001439: The Institute for Translational Sciences · The University of Utah — UL1TR002538: Uhealth Center for Clinical and Translational Science · Tufts Medical Center — UL1TR002544: Tufts Clinical and Translational Science Institute · Tulane University — UL1TR003096: Center for Clinical and Translational Science · The Queens Medical Center — None (Voluntary) · University Medical Center New Orleans — U54GM104940: Louisiana Clinical and Translational Science (LA CaTS) Center · University of Alabama at Birmingham — UL1TR003096: Center for Clinical and Translational Science · University of Arkansas for Medical Sciences — UL1TR003107: UAMS Translational Research Institute · University of Cincinnati — UL1TR001425: Center for Clinical and Translational Science and Training · University of Colorado Denver, Anschutz Medical Campus — UL1TR002535: Colorado Clinical and Translational Sciences Institute · University of Illinois at Chicago — UL1TR002003: UIC Center for Clinical and Translational Science · University of Kansas Medical Center — UL1TR002366: Frontiers: University of Kansas Clinical and Translational Science Institute · University of Kentucky — UL1TR001998: UK Center for Clinical and Translational Science · University of Massachusetts Medical School Worcester — UL1TR001453: The UMass Center for Clinical and Translational Science (UMCCTS) · University Medical Center of Southern Nevada — None (voluntary) · University of Minnesota — UL1TR002494: Clinical and Translational Science Institute · University of Mississippi Medical Center — U54GM115428: Mississippi Center for Clinical and Translational Research (CCTR) · University of Nebraska Medical Center — U54GM115458: Great Plains IDeA-Clinical & Translational Research · University of North Carolina at Chapel Hill — UL1TR002489: North Carolina Translational and Clinical Science Institute · University of Oklahoma Health Sciences Center — U54GM104938: Oklahoma Clinical and Translational Science Institute (OCTSI) · University of Pittsburgh — UL1TR001857: The Clinical and Translational Science Institute (CTSI) · University of Pennsylvania — UL1TR001878: Institute for Translational Medicine and Therapeutics · University of Rochester — UL1TR002001: UR Clinical & Translational Science Institute · University of Southern California — UL1TR001855: The Southern California Clinical and Translational Science Institute (SC CTSI) · University of Vermont — U54GM115516: Northern New England Clinical & Translational Research (NNE-CTR) Network · University of Virginia — UL1TR003015: iTHRIV Integrated Translational health Research Institute of Virginia · University of Washington — UL1TR002319: Institute of Translational Health Sciences · University of Wisconsin-Madison — UL1TR002373: UW Institute for Clinical and Translational Research · Vanderbilt University Medical Center — UL1TR002243: Vanderbilt Institute for Clinical and Translational Research · Virginia Commonwealth University — UL1TR002649: C. Kenneth and Dianne Wright Center for Clinical and Translational Research · Wake Forest University Health Sciences — UL1TR001420: Wake Forest Clinical and Translational Science Institute · Washington University in St. Louis — UL1TR002345: Institute of Clinical and Translational Sciences · Weill Medical College of Cornell University — UL1TR002384: Weill Cornell Medicine Clinical and Translational Science Center · West Virginia University — U54GM104942: West Virginia Clinical and Translational Science Institute (WVCTSI)· Submitted: Icahn School of Medicine at Mount Sinai — UL1TR001433: ConduITS Institute for Translational Sciences · The University of Texas Health Science Center at Tyler — UL1TR003167: Center for Clinical and Translational Sciences (CCTS) · University of California, Davis — UL1TR001860: UCDavis Health Clinical and Translational Science Center · University of California, Irvine — UL1TR001414: The UC Irvine Institute for Clinical and Translational Science (ICTS) · University of California, Los Angeles — UL1TR001881: UCLA Clinical Translational Science Institute · University of California, San Diego — UL1TR001442: Altman Clinical and Translational Research Institute · University of California, San Francisco — UL1TR001872: UCSF Clinical and Translational Science Institute· NYU Langone Health Clinical Science Core, Data Resource Core, and PASC Biorepository Core — OTA-21-015A: Post-Acute Sequelae of SARS-CoV-2 Infection Initiative (RECOVER)

Pending: Arkansas Children’s Hospital — UL1TR003107: UAMS Translational Research Institute · Baylor College of Medicine — None (Voluntary) · Children’s Hospital of Philadelphia — UL1TR001878: Institute for Translational Medicine and Therapeutics · Cincinnati Children’s Hospital Medical Center — UL1TR001425: Center for Clinical and Translational Science and Training · Emory University — UL1TR002378: Georgia Clinical and Translational Science Alliance · HonorHealth — None (Voluntary) · Loyola University Chicago — UL1TR002389: The Institute for Translational Medicine (ITM) · Medical College of Wisconsin — UL1TR001436: Clinical and Translational Science Institute of Southeast Wisconsin · MedStar Health Research Institute — None (Voluntary) · Georgetown University — UL1TR001409: The Georgetown-Howard Universities Center for Clinical and Translational Science (GHUCCTS) · MetroHealth — None (Voluntary) · Montana State University — U54GM115371: American Indian/Alaska Native CTR · NYU Langone Medical Center — UL1TR001445: Langone Health’s Clinical and Translational Science Institute · Ochsner Medical Center — U54GM104940: Louisiana Clinical and Translational Science (LA CaTS) Center · Regenstrief Institute — UL1TR002529: Indiana Clinical and Translational Science Institute · Sanford Research — None (Voluntary) · Stanford University — UL1TR003142: Spectrum: The Stanford Center for Clinical and Translational Research and Education · The Rockefeller University — UL1TR001866: Center for Clinical and Translational Science · The Scripps Research Institute — UL1TR002550: Scripps Research Translational Institute · University of Florida — UL1TR001427: UF Clinical and Translational Science Institute · University of New Mexico Health Sciences Center — UL1TR001449: University of New Mexico Clinical and Translational Science Center · University of Texas Health Science Center at San Antonio — UL1TR002645: Institute for Integration of Medicine and Science · Yale New Haven Hospital — UL1TR001863: Yale Center for Clinical Investigation

## References

[1] Long COVID or Post-COVID Conditions. U.S. Department of Health and Human Services, CDC (2021). https://www.cdc.gov/coronavirus/2019-ncov/long-term-effects/

[2] New ICD-10-CM code for Post-COVID Conditions, following the 2019 Novel Coronavirus (COVID-19). U.S. Department of Health and Human Services, CDC (2021). https://www.cdc.gov/nchs/data/icd/announcement-new-icd-code-for-post-covid-condition-april-2022-final.pdf

[3] Haendel, M.A., Chute, C.G., Bennett, T.D., Eichmann, D.A., Guinney, J., Kibbe, W.A., Payne, P.R., Pfaff, E.R., Robinson, P.N., Saltz, J.H., et al.: The national covid cohort collaborative (n3c): rationale, design, infrastructure, and deployment. Journal of the American Medical Informatics Association 28(3), 427–443 (2021)

[4] Hill, E.L., Mehta, H.B., Sharma, S., Mane, K., Xie, C., Cathey, E., Loomba, J., Russell, S., Spratt, H., DeWitt, P.E., et al.: Risk factors associated with postacute sequelae of sars-cov-2 in an ehr cohort: A national covid cohort collaborative (n3c) analysis as part of the nih recover program. medRxiv, 2022–08 (2022)

[5] Pfaff, E.R., Madlock-Brown, C., Baratta, J.M., Bhatia, A., Davis, H., Girvin, A., Hill, E., Kelly, E., Kostka, K., Loomba, J., et al.: Coding long covid: characterizing a new disease through an icd-10 lens. BMC medicine 21(1), 58 (2023)

[6] Reese, J.T., Blau, H., Casiraghi, E., Bergquist, T., Loomba, J.J., Callahan, T.J., Laraway, B., Antonescu, C., Coleman, B., Gargano, M., et al.: Generalisable long covid subtypes: findings from the nih n3c and recover programmes. EBioMedicine 87 (2023)

[7] Jiang, S., Loomba, J., Sharma, S., Brown, D.: Vital measurements of hospitalized covid-19 patients as a predictor of long covid: An ehr-based cohort study from the recover program in n3c. In: 2022 IEEE International Conference on Bioinformatics and Biomedicine (BIBM), pp. 3023–3030 (2022). IEEE

[8] Jager, K.J., Tripepi, G., Chesnaye, N.C., Dekker, F.W., Zoccali, C., Stel, V.S.: Where to look for the most frequent biases? Nephrology 25(6), 435–441 (2020)

[9] Austin, P.C., Lee, D.S., Fine, J.P.: Introduction to the analysis of survival data in the presence of competing risks. Circulation 133(6), 601–609 (2016)

[10] Schuster, N.A., Hoogendijk, E.O., Kok, A.A., Twisk, J.W., Heymans, M.W.: Ignoring competing events in the analysis of survival data may lead to biased results: a nonmathematical illustration of competing risk analysis. Journal of clinical epidemiology 122, 42–48 (2020)

[11] Sousa, G., Garces, T., Cestari, V., Florêncio, R., Moreira, T., Pereira, M.: Mortality and survival of covid-19. Epidemiology & Infection 148, 123 (2020)

[12] Neville, T.H., Hays, R.D., Tseng, C.-H., Gonzalez, C.A., Chen, L., Hong, A., Yamamoto, M., Santoso, L., Kung, A., Schwab, K., et al.: Survival after severe covid-19: long-term outcomes of patients admitted to an intensive care unit. Journal of intensive care medicine 37(8), 1019–1028 (2022)

[13] Long, J.D., Strohbehn, I., Sawtell, R., Bhattacharyya, R., Sise, M.E.: Covid-19 survival and its impact on chronic kidney disease. Translational Research 241, 70–82 (2022)

[14] Ibrahim, J.G., Chen, M.-H., Sinha, D., Ibrahim, J., Chen, M.: Bayesian survival analysis 2 (2001)

[15] Leung, K.-M., Elashoff, R.M., Afifi, A.A.: Censoring issues in survival analysis. Annual review of public health 18(1), 83–104 (1997)

[16] Joarder, A., Krishna, H., Kundu, D.: Inferences on weibull parameters with conventional type-i censoring. Computational statistics & data analysis 55(1), 1–11 (2011)

[17] Dutta, S., Dey, S., Kayal, S.: Bayesian survival analysis of logistic exponential distribution for adaptive progressive type-ii censored data. Computational Statistics, 1–47 (2023)

[18] Betancourt, M.: A conceptual introduction to hamiltonian monte carlo. arXiv preprint arXiv:1701.02434 (2017)

[19] Hoffman, M.D., Gelman, A., et al.: The no-u-turn sampler: adaptively setting path lengths in hamiltonian monte carlo. J. Mach. Learn. Res. 15(1), 1593–1623 (2014)

[20] Patil, A., Huard, D., Fonnesbeck, C.J.: Pymc: Bayesian stochastic modelling in python. Journal of statistical software 35(4), 1 (2010)

[21] Stuart, E.A., King, G., Imai, K., Ho, D.: Matchit: nonparametric preprocessing for parametric causal inference. Journal of statistical software (2011)

[22] Roffman, C., Buchanan, J., Allison, G.: Charlson comorbidities index. Journal of physiotherapy 62(3) (2016)

[23] OMOP Common Data Model. Observational Health Data Sciences and Informatics. https://ohdsi.github.io/CommonDataModel/

[24] N3C Privacy-Preserving Record Linkage. The National COVID Cohort Collaborative (N3C). https://covid.cd2h.org/PPRL/

[25] Adult BMI. U.S. Department of Health and Human Services, CDC. https://www.cdc.gov/healthyweight/assessing/bmi/adultbmi/index.html

[26] Rich, J.T., Neely, J.G., Paniello, R.C., Voelker, C.C., Nussenbaum, B., Wang, E.W.: A practical guide to understanding kaplan-meier curves. Otolaryngology—Head and Neck Surgery 143(3), 331–336 (2010)

[27] Bouman, P., Dukic, V., Meng, X.-L.: A bayesian multiresolution hazard model with application to an aids reporting delay study. Statistica Sinica, 325–357 (2005)

[28] Dukić, V., Dignam, J.: Bayesian hierarchical multiresolution hazard model for the study of time-dependent failure patterns in early stage breast cancer. Bayesian analysis (Online) 2(3), 591 (2007)

[29] Sinha, D., Chen, M.-H., Ghosh, S.K.: Bayesian analysis and model selection for interval-censored survival data. Biometrics 55(2), 585–590 (1999)

[30] Dutta, S., Dey, S., Kayal, S.: Bayesian survival analysis of logistic exponential distribution for adaptive progressive type-ii censored data. Computational Statistics 39(4), 2109–2155 (2024)

